# COFFEE CONSUMPTION AND PREVALENCE OF INSOMNIA DISORDER IN MEDICAL INTERNS AT THE AUTONOMOUS UNIVERSITY OF SANTO DOMINGO, DOMINICAN REPUBLIC, SEPTEMBER 2022-MARCH 2023

**DOI:** 10.1101/2024.11.18.24317358

**Authors:** Andy Bienvenido Ogando Montero, Inomar Nuñez Figuereo

**Affiliations:** Autonomous University of Santo Domingo INSIS-UASD, National District, Dominican Republic

**Keywords:** Insomnia, coffee consumption, medical interns, sleep disorders, Dominican Republic

## Abstract

This study aims to investigate the relationship between insomnia and coffee consumption among medical interns at the Autonomous University of Santo Domingo, Dominican Republic. The research stems from the observation of high coffee consumption among healthcare personnel, particularly those exposed to night shifts and conditions that impact sleep quality. A cross-sectional, descriptive, and quantitative study was conducted with a sample of 299 interns, divided into groups based on the presence or absence of insomnia symptoms according to DSM-V criteria. The results indicate a significant relationship between coffee consumption and the prevalence of insomnia, with higher incidence among interns who consume coffee daily. This study provides evidence on how coffee consumption, within the Dominican cultural context and the demands of the medical environment, can affect sleep quality among medical interns. These findings highlight the need to consider caffeine consumption as a potential factor in the assessment and management of sleep disorders within this group of healthcare professionals.

## Introduction

This study aims to shed scientific light on observable behaviors among our colleagues in the Faculty of Health Sciences. Here, we intend to determine the relationship between insomnia and coffee consumption.

In the Dominican Republic, drinking coffee is a deeply rooted tradition. Unlike other coffee-producing countries, domestic coffee consumption in this country is significant. National coffee demand was estimated at 2.81 kg of ground coffee per person in 2021 (Jiménez et al., 2007).

Healthcare personnel are also part of the general population and, therefore, are coffee consumers. Moreover, this group is often exposed to additional factors, such as night shifts, which cause not only tiredness but exhaustion, as short sleep periods are impossible, and the fatigue generated by staying awake for long hours at night is hard to compensate. In this study, however, we will focus exclusively on coffee consumption as a variable to assess its relationship with the prevalence of insomnia.

## Methods

This is a cross-sectional, descriptive, quantitative study. The study population consisted of medical interns at the UASD (Universidad Autónoma de Santo Domingo) completing their rotational internship during the periods of September 2022 and March 2023.

We used simple random sampling to select the participants, dividing them into three groups:

1. Interns with insomnia.
2. Interns meeting diagnostic criteria for insomnia according to the DSM-5 (The Diagnostic and Statistical Manual of Mental Disorders, 5th Edition).
3. Interns without insomnia.

### Survey Instrument

Data were collected using a 15-question structured survey, which was designed to classify participants into the three aforementioned categories and further separate them by gender. The survey included the following topics:

- Topic 1: age, gender.
- Topic 2: Sleep habits and symptoms, incorporating validated criteria from the DSM-5 to identify insomnia symptoms and diagnose insomnia.
- Topic 3: Coffee consumption habits, including:
  - Frequency of coffee intake.
  - Number of cups consumed per day.
  - Time of day when coffee is consumed.

### Study Population and Sampling

The target population consisted of 1,356 medical interns in the September 2022/2023 and March 2023/2024 rotations in hospitals affiliated with the UASD School of Medicine. Using Epi Info 7.2, we calculated a required sample size of 299 participants to achieve a 95% confidence interval and a margin of error of 5%. Participants were selected randomly from the complete list of interns provided by the university administration.

### Data Collection Process

Participants were invited to complete the survey via an online platform, ensuring anonymity and confidentiality. The survey system was programmed to classify participants automatically into one of the three groups based on their responses:

- Interns with insomnia: Those meeting the DSM-5 diagnostic criteria.
- Interns meeting partial criteria: Those with symptoms consistent with insomnia but insufficient to meet the full diagnostic criteria.
- Interns without insomnia: Those reporting no symptoms of insomnia.

### Ethical Considerations

The study protocol was approved by the Ethics Committee of the UASD School of Medicine. Informed consent was obtained from all participants before administering the survey.

### Data Analysis

Data were analyzed using descriptive, inferential, and correlational statistics. Descriptive statistics summarized the prevalence of insomnia and coffee consumption by gender and category (Insomnia, With Criteria, Without Insomnia). Inferential statistics were used to evaluate differences between groups, assessing statistical significance at a 95% confidence level.

A correlation analysis was performed to explore the relationship between daily coffee consumption and the prevalence of insomnia across the three categories (Insomnia, With Criteria, Without Insomnia). Pearson’s correlation coefficient was calculated to measure the strength and direction of the association, and significance was determined at the p < 0.05 level. The results were visualized to illustrate patterns by gender and category, highlighting differences in coffee consumption and insomnia prevalence.

This comprehensive approach allowed for a deeper understanding of the factors contributing to insomnia and their association with coffee consumption, providing insights into gender-specific patterns and their implications.

## Supplementary Materials

The complete survey instrument, including all sections and questions, has been provided as a supplemental file to allow qualified readers to follow and replicate.

### Questionary

1. **Are you diagnosed with an insomnia disorder?** Responses: Yes / No.
2. **Gender:** Responses: Male / Female.
3. **Do you consume coffee?** Responses: Yes / No.
4. **Do you drink coffee?** Responses: Yes / No.
5. **How many cups of coffee do you drink daily?** Responses: I don’t drink coffee / 1 / 2 / 3 / 4 or more.
6. **How many cups of coffee do you consume per day?** Responses: I don’t drink coffee / 1 / 2 / 3 / 4 or more.
7. **How many cups do you drink per day?** Responses: I don’t drink coffee / 1 / 2 / 3 / 4 or more.
8. **How many cups of coffee do you drink in a day?** Responses: I don’t drink coffee / 1 / 2 / 3 / 4 or more. **Gender:** Responses: Male / Female.
9. **Do you drink coffee?** Responses: Yes / No.
10. **Do you drink coffee?** Responses: Yes / No.
11. **How many cups of coffee do you consume daily?** Responses: I don’t drink coffee / 1 / 2 / 3 / 4 or more.
12. **How many cups of coffee do you drink per day?** Responses: I don’t drink coffee / 1 / 2 / 3 / 4 or more.
13. **Have you experienced any of the following events?** Responses: Difficulty initiating sleep. Difficulty maintaining sleep. Poor sleep quality. None.
14. **How many cups of coffee do you drink per day?** Responses: I don’t drink coffee / 1 / 2 / 3 / 4 or more.

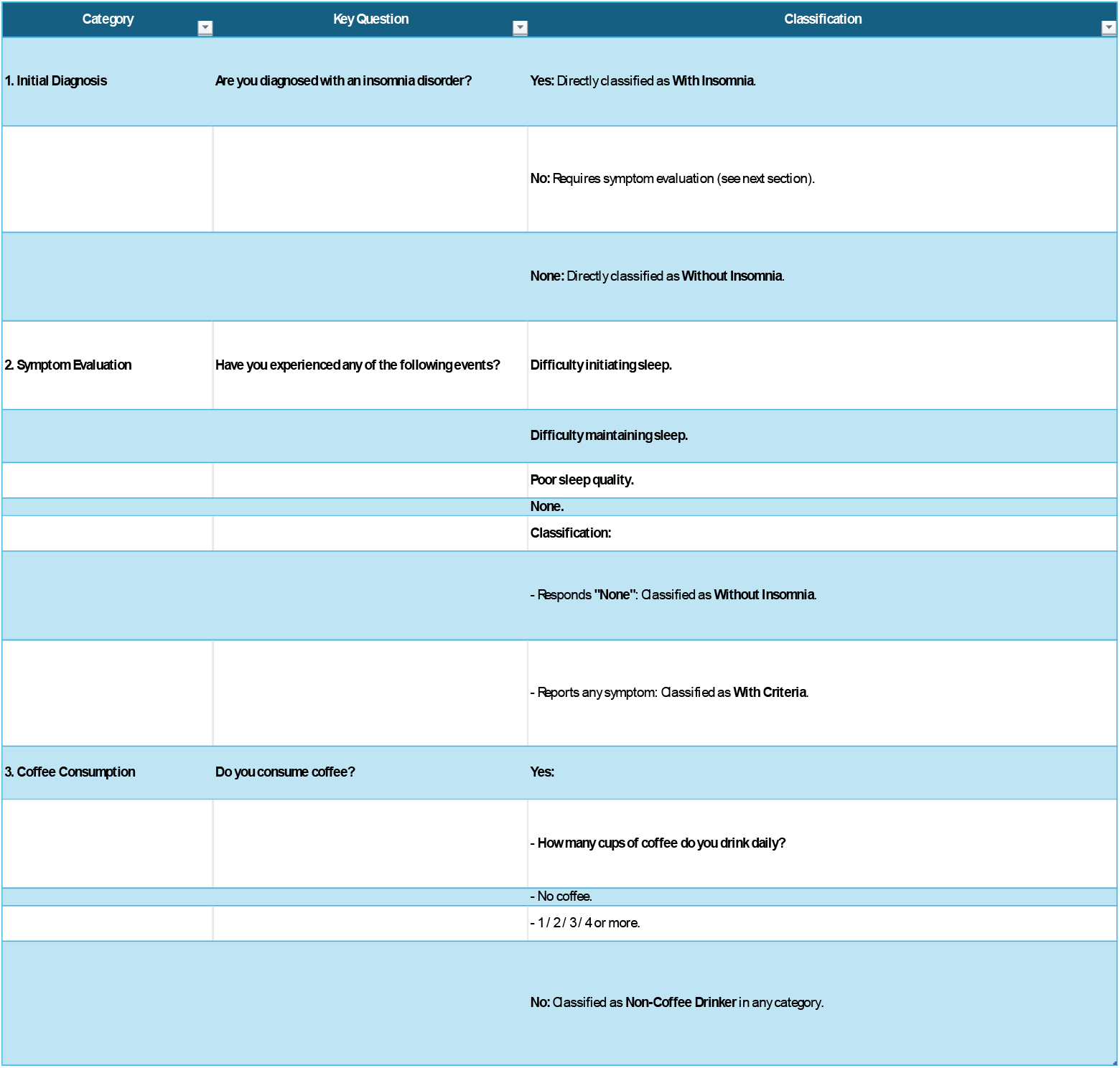

| Category | Key Question | Classification |
| --- | --- | --- |
| 1. Initial Diagnosis | Are you diagnosed with an insomnia disorder? | Yes: Directly classified as **With Insomnia**. |
|  |  | No: Requires symptom evaluation (see next section). |
|  |  | None: Directly classified as **Without Insomnia**. |
| 2. Symptom Evaluation | Have you experienced any of the following events? | Difficulty initiating sleep. |
|  |  | Difficulty maintaining sleep. |
|  |  | Poor sleep quality. |
|  |  | None. |
|  |  | Classification: |
|  |  | - Responds "**None**": Classified as **Without Insomnia**. |
|  |  | - Reports any symptom: Classified as **With Criteria**. |
| 3. Coffee Consumption | Do you consume coffee? | Yes: |
|  |  | - How many cups of coffee do you drink daily? |
|  |  | - No coffee. |
|  |  | - 1 / 2 / 3 / 4 or more. |
|  |  | No: Classified as **Non-Coffee Drinker** in any category. |

## Results

The findings of this research indicate a relationship between coffee consumption and the prevalence of insomnia and/or symptoms thereof (criteria).

A total of 299 interns were surveyed, consisting of 61 men and 238 women (Table #1).

**Table 1.**
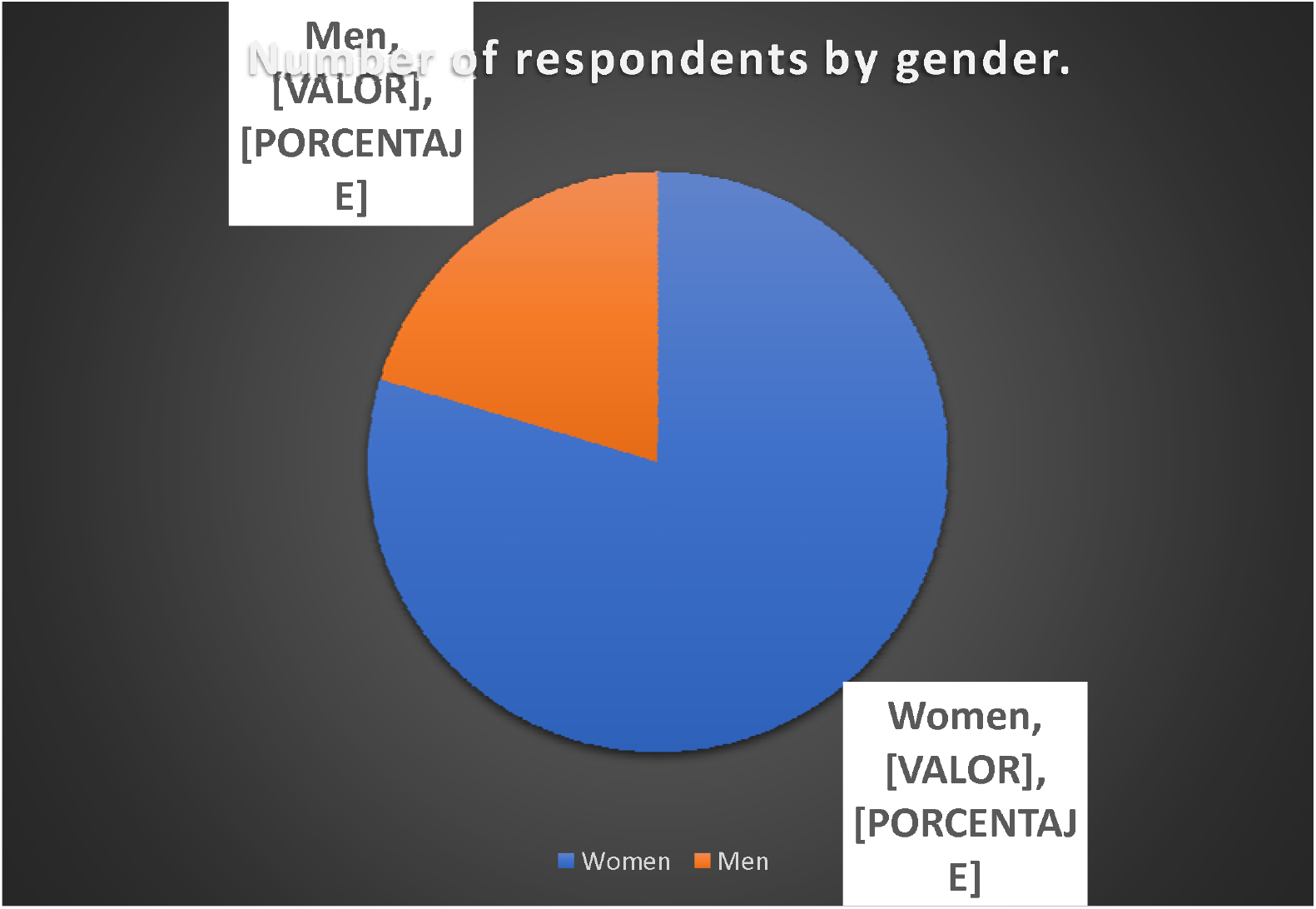

- For the 61 men, the following data were obtained: 17 (28%) without insomnia, 35 (57%) with criteria, and 9 (15%) with insomnia (Table #2).
- For the 238 women, the following data were obtained: 58 (24%) without insomnia, 166 (70%) with criteria, and 14 (6%) with insomnia (Table #3).
- The number of interns who reported coffee consumption was higher in both sexes and across all categories than those who did not consume coffee. Among the population presenting criteria and/or insomnia, totaling 226 individuals, 144 (63%) consumed coffee, while 82 (36%) did not. This indicates a higher prevalence of insomnia among coffee consumers (Table #4).
- There is a trend toward higher daily coffee intake among those with insomnia (1.8 cups for men; 2.1 cups for women) and those with diagnostic criteria (1.9 cups for men; 1.7 cups for women) than among those without insomnia (1.4 cups for men; 1.2 cups for women) (Table #5).
- Among women, there is a directly proportional relationship between coffee consumption and insomnia prevalence, with a higher number of cups corresponding to a higher prevalence of criteria and/or insomnia (1.7 cups; 2.1 cups, respectively) (Table #5).
- An atypical relationship was observed in men, where those meeting criteria consumed an average of 1.9 cups per day, higher than those with insomnia (1.8 cups). This may be due to other factors, including diet-hygienic or diet-therapeutic measures that might require lower coffee consumption among those diagnosed. While we cannot confirm this, future studies could explore this hypothesis. Nonetheless, the mean coffee consumption across all categories and in both sexes was lower for those without insomnia.
- We determined that there is no significant relationship between coffee consumption and gender, as percentages were very similar in both groups (Table #5).

**Table 2.**
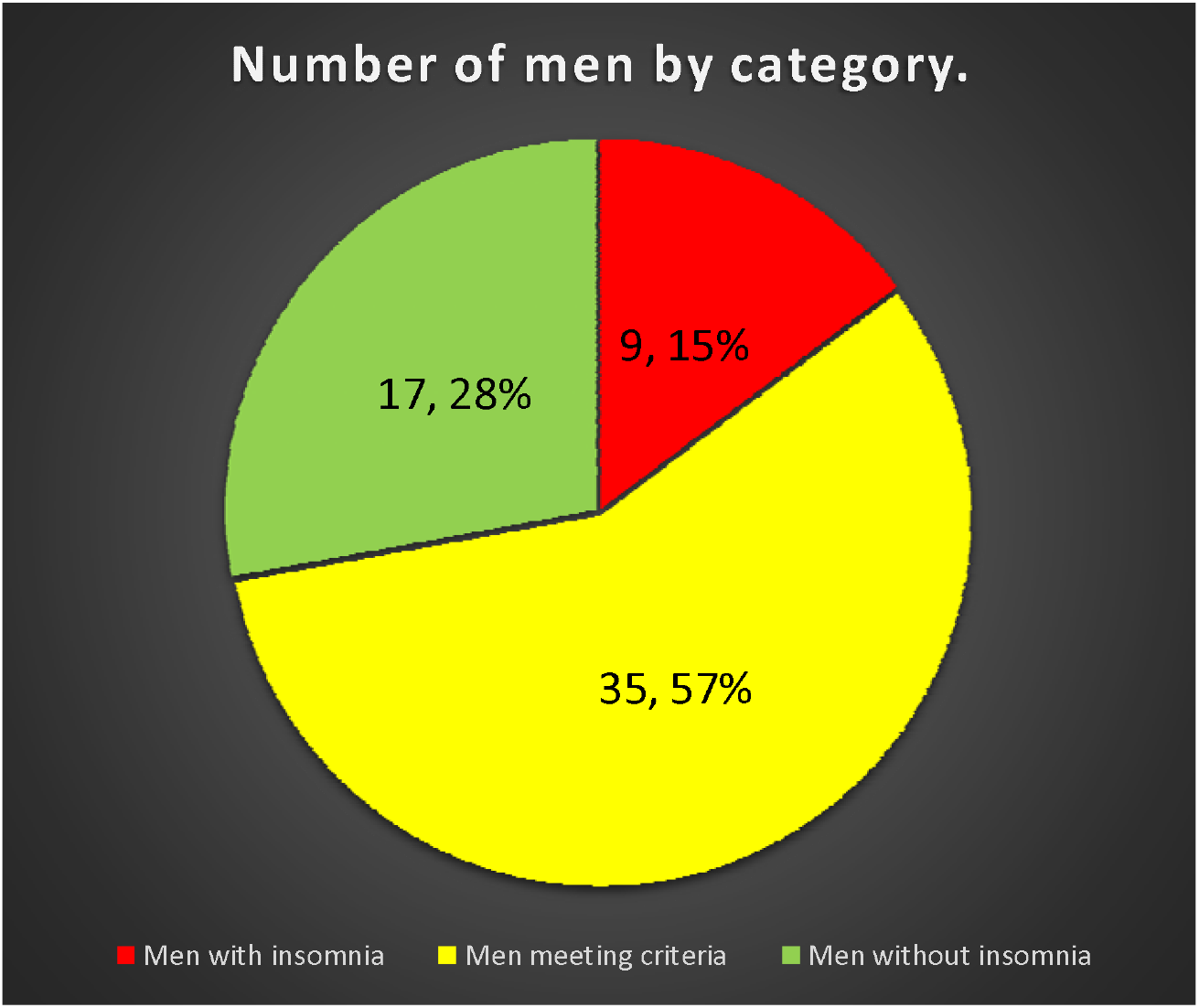

**Table 3.**
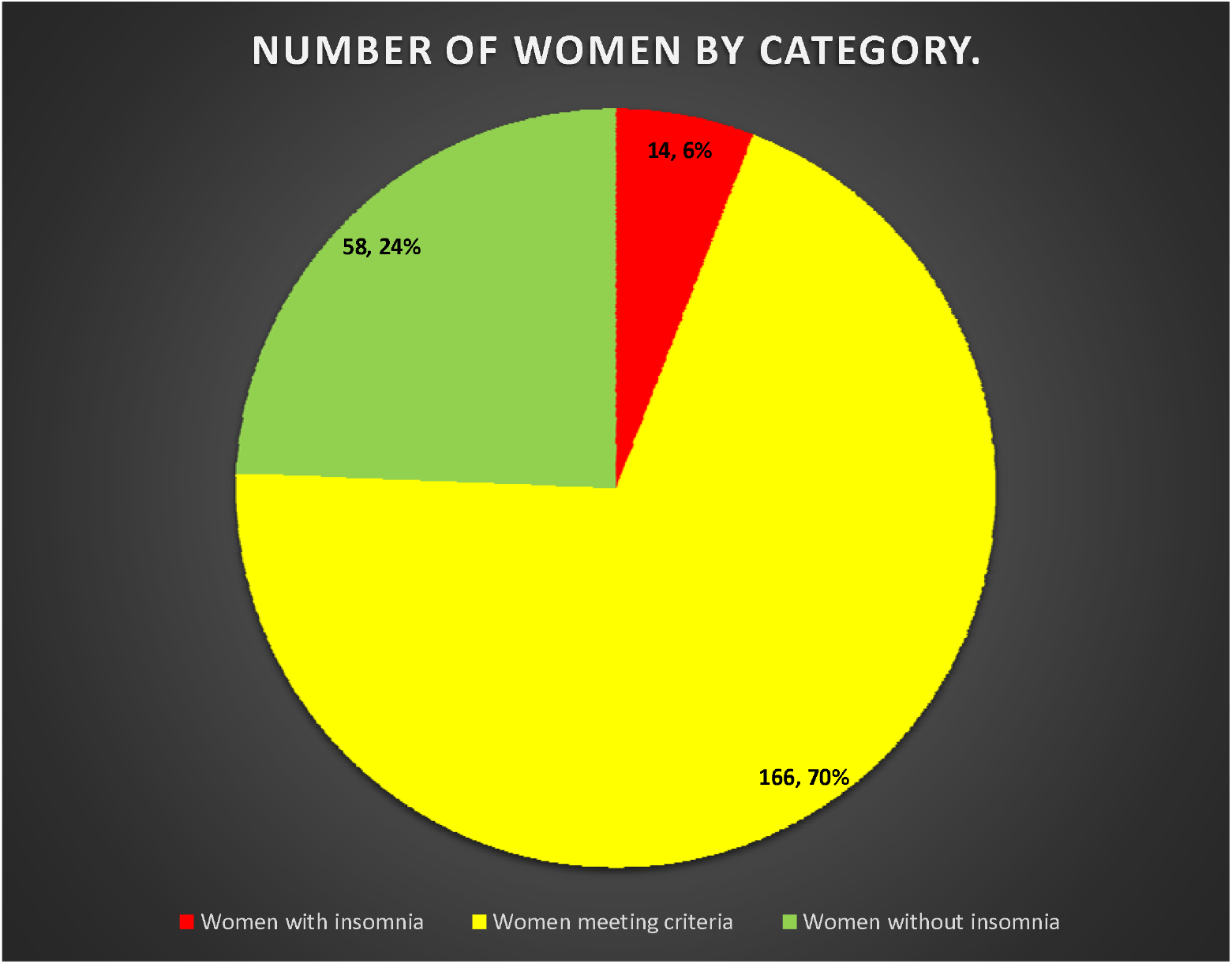

**Table 4.**
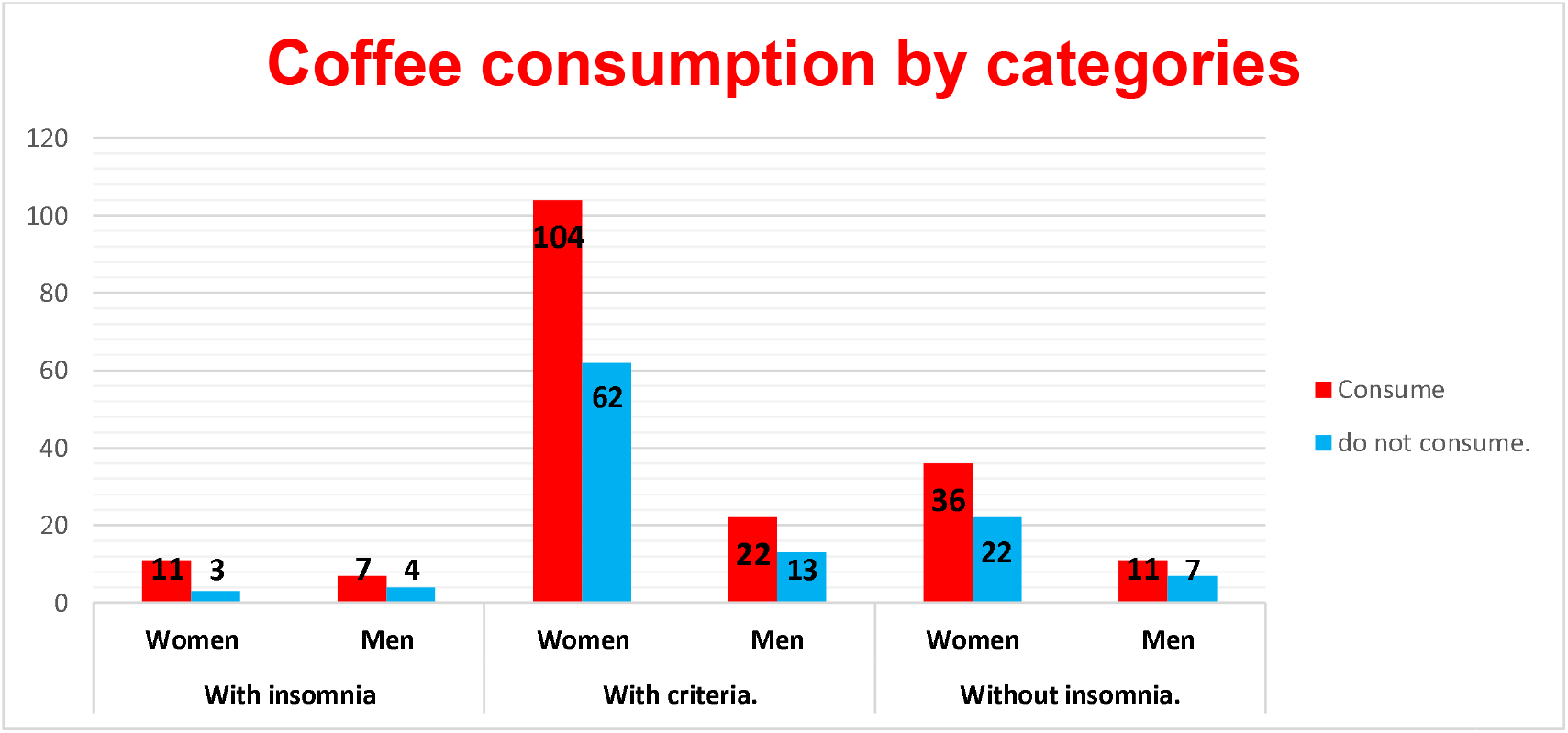

**Table 5.**
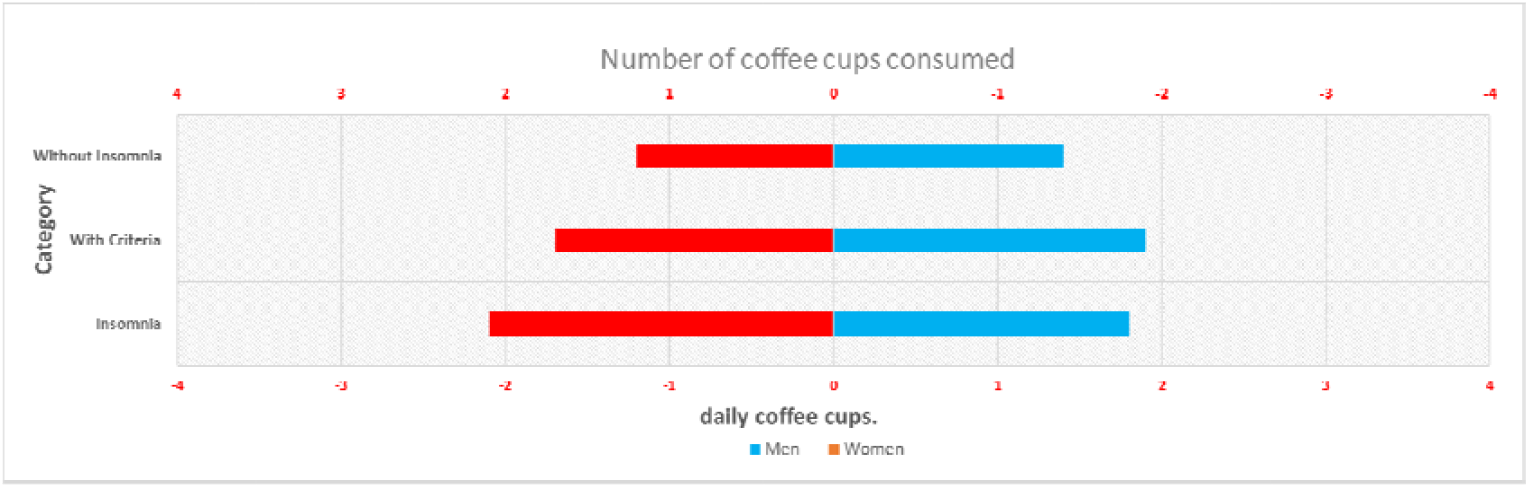

## Discussion

The consumption of coffee among medical interns was higher than described in other studies, although the relationship between insomnia and clinical symptoms of insomnia is similar, as shown in the study by Abdelmoaty Goweda R, Hassan-Hussein A. in 2020, in their article *“Prevalence of sleep disorders among medical students of Umm Al-Qura University, Makkah, Kingdom of Saudi Arabia*.*”* In this study, 73.8% of participants had sleep disorders, a percentage very similar to our study, which reported 74% on average for both sexes. Similarly, the results of the study *“Use of energy drinks and symptoms of insomnia in medical students of a Peruvian university”* by Mendoza L. Marco G, demonstrate the prevalence of insomnia symptoms in the studied sample, which consumed energy drinks (high levels of caffeine). These findings align with the data shown in this study.

Based on the presented results, we define the relationship between coffee consumption and the prevalence of insomnia, which should be considered as an associated factor.

## Data Availability

All data produced in the present work are contained in the manuscript

